# Impact of mitigating interventions and temperature on the instantaneous reproduction number in the COVID-19 epidemic among 30 US metropolitan areas

**DOI:** 10.1101/2020.04.26.20081083

**Authors:** Xinhua Yu

**Affiliations:** Division of Epidemiology, Biostatistics and Environmental Health, School of Public Health, University of Memphis

**Keywords:** virus transmissibility, reproduction number, mitigating intervention, temperature, epidemic, COVID-19

## Abstract

**Background:** After more than four months into the coronavirus disease (COVID-19) epidemic, over 347,500 people had died worldwide. The current study aims to evaluate how mitigating interventions affected the epidemic process in the 30 largest metropolitan areas in the US and whether temperature played a role in the epidemic process.

**Methods:** Publicly available data for the time series of COVID-19 cases and deaths and weather were analyzed at the metropolitan level. The time-varying reproductive numbers (R_t_) based on retrospective moving average were used to explore the trends. Student t tests were used to compare temperature and peak R_t_ cross-sectionally.

**Results:** We found that virus transmissibility, measured by instantaneous reproduction number (R_t_), had declined since the end of March for all areas and almost all of them reached a R_t_ of 1 or below after April 15, 2020. However, the R_t_s remained around 1 for most areas since then and some small and short rebounds were presented in some areas, suggesting a persistent epidemic in those areas. The timing of the main decline was concurrent with the implementation of mitigating interventions. Cities with warm temperature also tended to have a lower peak R_t_ than that of cities with cold temperature. However, large geographic variations existed.

**Conclusions:** Aggressive interventions might have mitigated the current epidemic of COVID-19, while temperature might have some weak effects on the virus transmission. We may need to prepare for a possible return of the coronavirus outbreak.

## Introduction

The coronavirus disease (COVID-19) pandemic caused by the novel severe acute respiratory syndrome (SARS) associated coronavirus (SARS-CoV2) infection [1] has not only affected more than 5.5 million people and caused over 347,500 deaths worldwide (https://coronavirus.jhu.edu/map.html, accessed May 26, 2020), but also induced significant anxiety among the public [2]. Many people raised concerns about whether the stringent interventions were over-reacted, and whether a second wave of outbreak was possible. In the 2003 SARS epidemic, the virus went away after June 2003 and never came back[3]. Will this happen to SARS-CoV2?

There are several major differences between 2003 SARS coronavirus and 2019 SARS CoV2 [3, 4]. The 2003 coronavirus had much higher virulence, resulting in higher hospitalizations and mortality rates than the 2019 coronavirus. The transmission of the 2003 coronavirus almost exclusively occurred among symptomatic cases[5], while the 2019 coronavirus can cause a large percent of asymptomatic cases who can also transmit virus [6, 7]. Furthermore, the 2019 coronavirus is also circulating in the southern hemisphere where the current temperature is warmer than that of northern hemisphere. By late 2020, it is possible the coronavirus may circulate back to the northern hemisphere, leading to a second wave of epidemic[8]. Therefore, it is important to empirically evaluate the effects of mitigating interventions and examine whether temperature may affect the virus transmissibility and virulence.

One key measure of virus transmissibility during an epidemic is effective reproduction number (R), the average number of secondary cases infected by a primary case [9–11]. Based on the susceptible-infectious-removed (SIR) model, the reproduction number can be conceptualized as (number of contacts)*(infectivity per contact)*(generation interval), where generation interval refers to the average duration between the time when a primary case becoming infectious and the time when secondary cases being infected[10]. Clearly, interventions such as social distancing, stay-at-home rule, school or office closures, and prohibiting large gatherings will reduce the number of contacts, thus reducing R. On the other hand, a lower virus infectivity can also reduce R, assuming the number of contacts remains unchanged. Therefore, exploring the changes of R over time and across different regions can shed new lights on the impact of interventions and environmental factors during the epidemic.

A few studies have used time varying R_t_ to explore the effects of intervention on the epidemic process[12–16]. For example, one recent study found significant effects of nonpharmaceutical interventions on the transmissibility of SARS-CoV2, measured by R_t_, in Hongkong[16]. However, few studies examined the impact of environmental factors on the COVID-19 epidemic. A few unpublished manuscripts have examined the association between temperature and COVID-19 case counts and found no or a negative but weak association[17–19]. However, their studies were based on case counts among different countries, which subjected to myriads of confounding effects due to different diagnostic criteria, availability of detection kits and reporting biases.

In this study, we will compare the magnitude and changes of time-varying (instantaneous) effective reproduction numbers (R_t_) among 30 largest metropolitan areas in the US. We hypothesize that stringent interventions are effective in curbing the epidemic, but temperature may also facilitate the decline of the epidemic in some regions.

## Methods

### Data source

We obtained daily COVID-19 cases and deaths at the US county level from the data repository provided by New York Times (https://github.com/nytimes/covid-19-data, accessed on May 26,2020). We further limited those counties to the 30 largest metropolitan areas (Table 1). All cases and deaths were summarized at the metropolitan level. The sizes of total population and people aged 65 or above for each metropolitan area were obtained from census bureau website. Information about stay-at-home rule for each state was scraped from popular news media. The historical daily average temperature was obtained from national climate data online (https://www.7.ncdc.noaa.gov/CDO), mostly based on temperature collected from stations at each metropolitan’s main airport.

**Table 1:** Characteristics and death rates of COVID-19 for 30 largest metropolitan areas in the US, as of May 25, 2020.

| **Metropolitan area** | **Total population** | **Age >=65** | **Cumulative cases** | **Cumulative deaths** | **Deaths per 100 Cases** | **Deaths per 100 old people** |
| --- | --- | --- | --- | --- | --- | --- |
| New York | 16,669,277 | 2,789,981 | 185,447 | 11,870 | 6.40 | 0.43 |
| Los Angeles | 10,039,107 | 1,493,190 | 46,018 | 2,116 | 4.60 | 0.14 |
| Washington DC - Baltimore | 9,360,001 | 1,370,637 | 64,441 | 2,572 | 3.99 | 0.19 |
| Houston | 7,066,141 | 865,112 | 15,306 | 331 | 2.16 | 0.04 |
| St. Francisco - St. Jose | 6,463,637 | 991,147 | 11,269 | 388 | 3.44 | 0.04 |
| Chicago | 6,021,020 | 950,164 | 87,730 | 4,012 | 4.57 | 0.42 |
| St Louis | 5,485,267 | 797,415 | 9,696 | 755 | 7.79 | 0.09 |
| Atlanta | 5,261,067 | 741,175 | 20,764 | 879 | 4.23 | 0.12 |
| Dallas-Fort Worth | 5,081,942 | 615,560 | 12,068 | 302 | 2.50 | 0.05 |
| Philadelphia | 3,815,431 | 691,016 | 44,325 | 2,772 | 6.25 | 0.40 |
| Minneapolis-St. Paul | 3,654,908 | 580,638 | 13,699 | 766 | 5.59 | 0.13 |
| Cleveland - Akron | 3,149,448 | 669,206 | 7,701 | 642 | 8.34 | 0.10 |
| Seattle | 3,074,865 | 444,891 | 11,130 | 687 | 6.17 | 0.15 |
| Boston | 2,979,288 | 529,944 | 43,788 | 2,842 | 6.49 | 0.54 |
| Denver | 2,967,239 | 419,589 | 15,250 | 908 | 5.95 | 0.22 |
| Miami - Fort Lauderdale-West Palm Beach | 2,716,940 | 467,586 | 17,040 | 633 | 3.71 | 0.14 |
| Charlotte | 2,675,243 | 424,892 | 5,949 | 165 | 2.77 | 0.04 |
| Orlando | 2,608,147 | 430,027 | 3,224 | 83 | 2.57 | 0.02 |
| Portland | 2,492,412 | 424,694 | 2,545 | 117 | 4.60 | 0.03 |
| Sacramento - Oakland | 2,363,730 | 419,255 | 1,762 | 88 | 4.99 | 0.02 |
| Pittsburg | 2,317,600 | 542,666 | 3,324 | 294 | 8.84 | 0.05 |
| Las Vegas | 2,266,715 | 358,821 | 6,182 | 331 | 5.35 | 0.09 |
| Cincinnati | 2,198,450 | 394,794 | 5,145 | 256 | 4.98 | 0.06 |
| Kansas | 2,157,990 | 368,576 | 4,096 | 170 | 4.15 | 0.05 |
| Columbus | 2,122,271 | 325,706 | 8,466 | 311 | 3.67 | 0.10 |
| Detroit | 2,116,944 | 387,897 | 21,114 | 2,460 | 11.65 | 0.63 |
| Indianapolis | 2,074,537 | 337,623 | 14,764 | 1,053 | 7.13 | 0.31 |
| Durham-Raleigh | 1,974,709 | 289,249 | 4,213 | 184 | 4.37 | 0.06 |
| Salt Lake City | 1,880,948 | 207,100 | 6,428 | 82 | 1.28 | 0.04 |
| Milwaukee | 1,575,179 | 288,873 | 7,314 | 316 | 4.32 | 0.11 |

### Reproduction number and serial interval

The time varying reproduction number (R_t_) was proposed by Cori A. et al. [11, 20]. This approach assumes the occurrence of secondary cases follows a Poisson distribution, conditioning on the time position in the whole infectious period of the primary case. The overall R_t_ at time t of the epidemic is the average number of secondary cases for all prior infected cases who are still infectious at a time window (t-s, t). The estimate is based on current secondary cases, not the future infections. Therefore, it can be viewed as instantaneous R_t_. To smooth the estimates, a weighted average of R_t_ over a sliding window is used (e.g., one week window). Consequently, we used a 7-day moving average of prior temperature when assessing the association between R_t_ and temperature.

The estimation of R_t_ also depends on the estimation of virus infectivity profile that is approximated from the distribution of generation interval. Since it is difficult to accurately estimate the generation interval, serial interval is often used in calculating R_t_ [11]. The serial interval refers to the average duration between symptom onset of a primary case and symptom onset of secondary cases. In this study, we assumed the serial interval had a gamma distribution with mean=4.7 days and standard deviation = 2.9 days [21, 22], similar to that of recent studies[15]. This serial interval was applied to all regions throughout the whole study period to ensure the compatibility of R_t_ across regions. However, different regions at different time might have different serial intervals due to interventions and other environmental and social factors.

We also performed sensitivity analysis with a shorter serial interval (mean = 3.95, standard deviation=4.75 [23] and a longer interval (mean = 7.5, standard deviation = 3.4) [4]. The patterns of R_t_ were similar, except for different estimated R_t_ values (average peak R_t_ 1-4 for shorter duration, and 2-9 for longer duration).

### Statistical analysis

Descriptive statistics and bivariate associations were reported. Student t-tests were used for comparisons. The sizes of total population and people aged 65 or older, and the percent of positive tests at each date were used for adjustment. R package EpiEstim was used [11] to estimate the instantaneous reproduction numbers over time. The association between temperature and R_t_ was explored cross-sectionally at the peak of R_t_ and also at some arbitral dates.

We adopted two time scales in the analysis. The first was calendar date to present the trends of reproduction numbers for all metropolitan areas, starting from the date with at least 10 total reported cases. Staggered entrances into the outbreak were preserved. The second scale was the time since the beginning of the outbreak, regardless what calendar date the outbreak happened. This was to compare the declining patterns of R_t_ across metropolitan areas. We also realigned the time scale from the peak of the outbreak. The first two weeks of R_t_ estimates were excluded, as the first week R_t_ were zeros, and the second week estimates were too variable due to small number of cases.

P value less than 0.05 was considered statistically significant. However, there were many statistical comparisons involved. Although we did not adjust for multiple comparisons, we were cautious about over interpretations and conducted statistical tests only between prior selected pairs (e.g., southern versus northern metropolitan areas).

### Ethics statement

In this study, the author has no financial and conflict of interest to disclose. The ethics approval was exempted for this study, as no human subjects were involved, and all data were publicly available. The statistical codes and data will be available online (https://tinyurl.com/ybhdcjqu).

## Results

The basic characteristics of metropolitan areas were presented in Table 1. All metropolitan areas had at least 1.5 million people in 2019 and over 1,000 confirmed cases. As of May 25, 2020, the reported case-fatality rates varied from 1.28 per 100 cases in Salt Lake City, UT to 11.65 per 100 cases in Detroit, MI. However, since there were large variations in case ascertainment criteria and availability of detection kits among different regions, comparing case-fatality rates was unreliable. Meanwhile, since about 80% of deaths occurred among elderly people[24], we compared the ratios of deaths to the size of elderly population among metropolitan areas. The ratios were generally lower in areas with warm weather (mean 0.09, range 0.02- to 0.22 per 100 elderly), and higher in areas with cold temperature (mean: 0.24, range 0.05 to 0.63 per 100 elderly) (p for difference = 0.007).

The trends of R_t_ for 30 metropolitan areas were shown in Figure 1a-1d, grouped by geographic locations and temperature conditions. Overall, the instantaneous R_t_s in all areas reached peaks or some stable points after two to three weeks, decreased significantly since the end of March, and most areas reached a R_t_ of 1 or less after April 15. However, some small and short rebounds were presented in some areas. This might be due to case reporting and detection issues, but could also indicate some true rebounds. In addition, the R_t_s remained around 1 for most areas, suggesting a persistent epidemic in those areas. It is of note that around the week of March 25, many schools were closed and many companies started offering employees working from home. The US government has issued COVID-19 coping guideline to all US citizens, and many states also issued stay-at-home rules (see Appendix Table 1).

**Figure 1a-1d:**
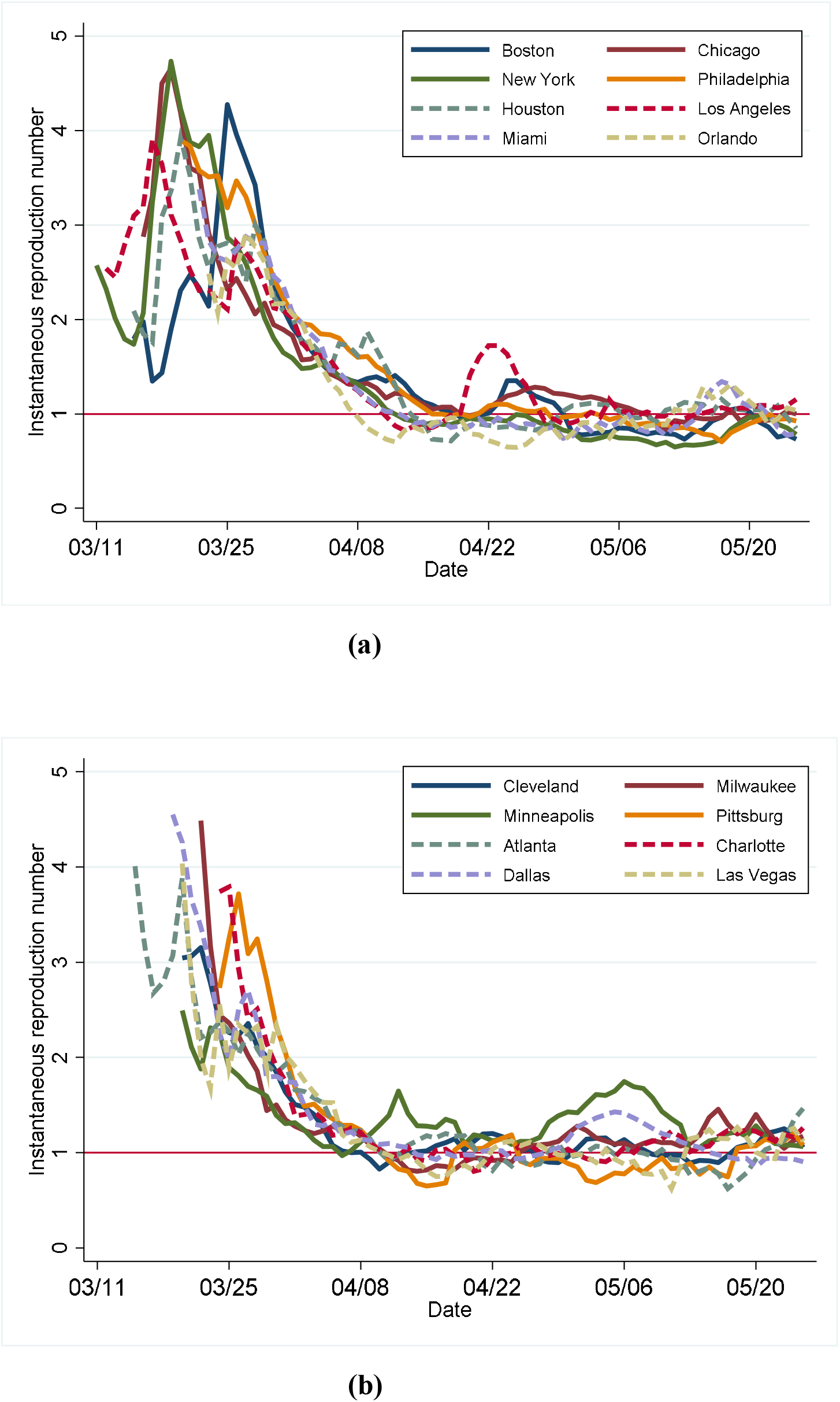
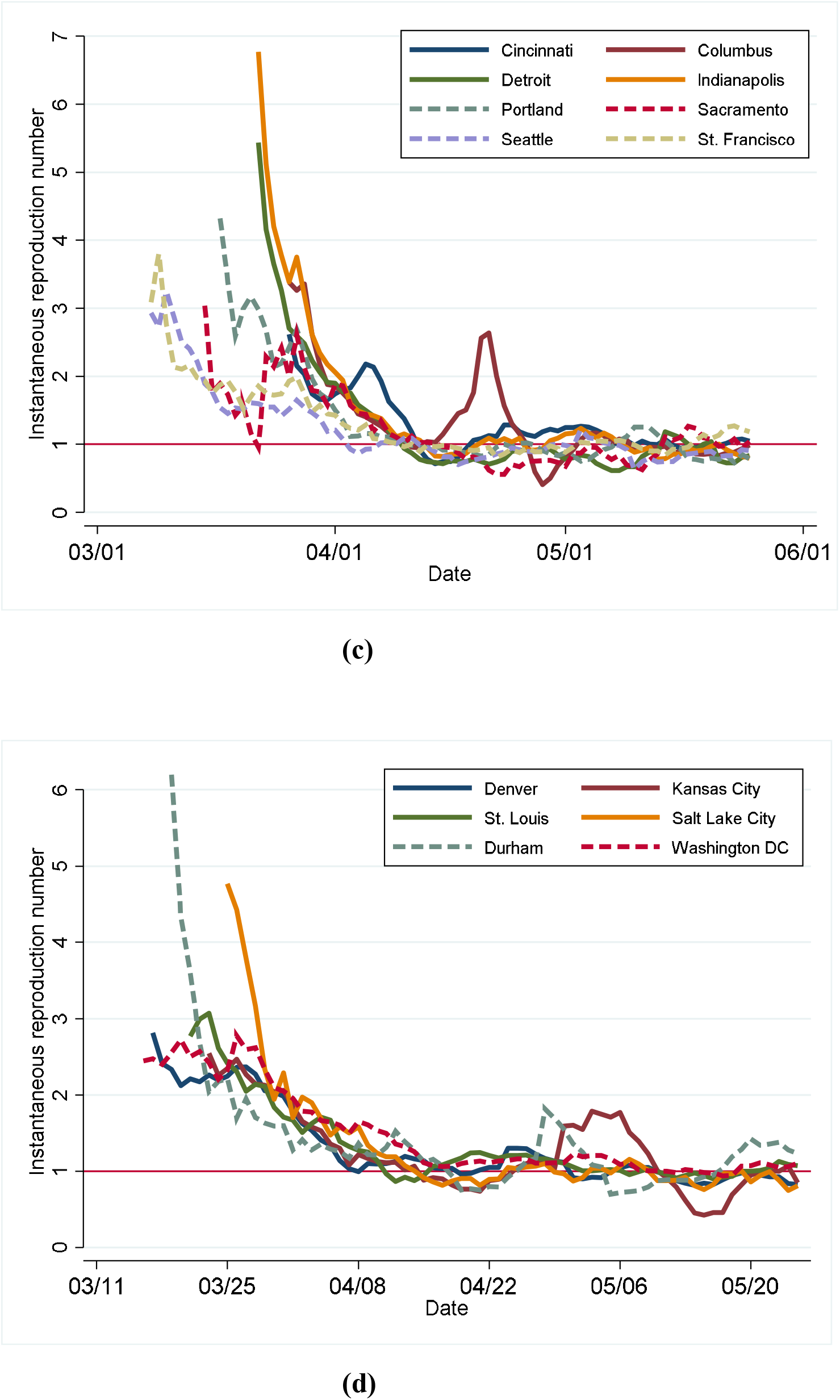
Time trends of instantaneous reproduction number for 30 US largest metropolitan areas.

Figure 1a compared R_t_ trends between typical northeastern cities and southern cities. Boston, Chicago, New York and Philadelphia started the epidemic earlier, had higher peak R_t_s than that of Miami, Orlando, Houston, and Los Angeles. After some initial increases (though peaked at different dates), all northern cities declined sharply after the mid-March. In addition, the trajectories of Houston and Los Angeles were similar with initial peaks at around March 18, somewhat decreased and then were stable around March 25. For Miami and Orland, the R_t_s were quite stable during the week of March 25, and declined sharply after about March 28. However, the slopes of decline, when aligned by the time since the peak R_t_, were similar except a few spikes in Houston and Los Angeles (Appendix Figure 1a).

On the other hand, the R_t_ curves were indistinguishable between upper midwestern cities and other southern cities (Figure 1b). Upper midwestern cities except Pittsburg all had earlier interventions in the mid-March (Appendix Table 1 for dates stay-at-home rules issued), while Minneapolis-St. Paul area had some rebounds over the course of epidemic. In addition, the west coastal cities had an early start of the epidemic, and R_t_ curves were less volatile than that of other cities during the study period (Figure 1c). The unusually high R_t_ in Salt Lake City in the early epidemic may be due to a small number of cases during that period (Figure 1d).

Furthermore, after realigning the starting time from their respective outbreak peaks for all metropolitan areas, the overall declining patterns were similar across all regions (Appendix Figure 1a-d).

To evaluate the association between R_t_ and temperature across regions, we compared the highest R_t_ (occurred after the first two weeks) among them (Figure 2), most cities had a peak R_t_ between 1 and 5. At the low temperature, peak R_t_s varied significantly. Cities with cold average temperature generally had higher R_t_s than those with warm temperature. However, upper midwestern cities such as Minneapolis-St. Paul, Milwaukee, and Columbus had much lower peak R_t_s than the rest of cities. The average peak of R_t_s in Boston, Chicago, New York, and Philadelphia were marginally (but not statistically significant) higher than that of Houston, Los Angeles, Orlando and Miami (average peak R_t_ 4.01 vs. 3.15, p = 0.07). In addition, we also arbitrarily examined the R_t_ patterns on the 15^th^ day after the outbreak and on March 24, 2020 when most interventions had not fully executed (Appendix Figure 2a and 2b). These cross-sectional analyses demonstrated similar patterns to that of peak R_t_.

**Figure 2:**
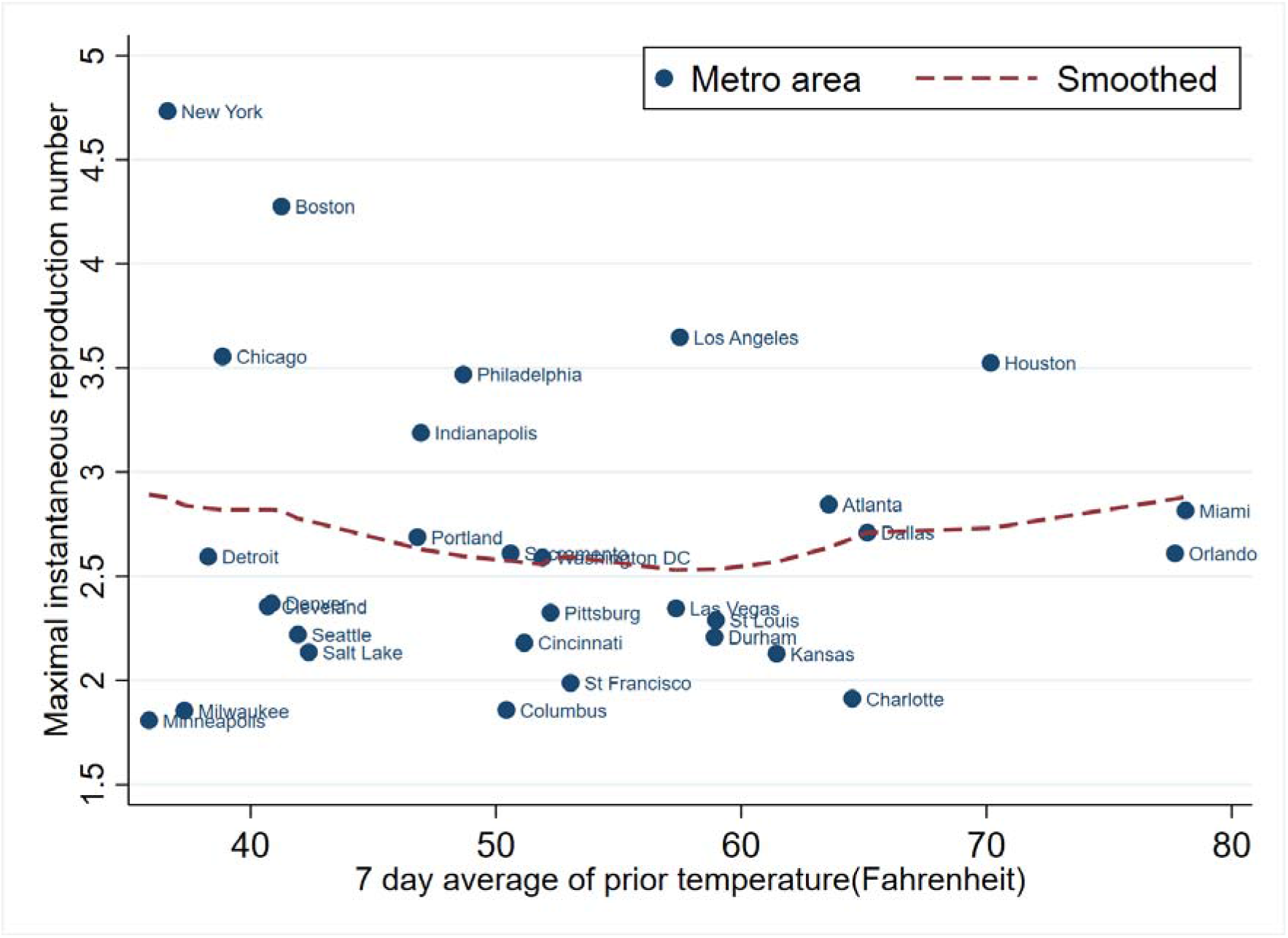
Association between maximal reproduction number and 7-day average temperature among US 30 metropolitan areas.

## Discussion

Overall, since the end of March, the instantaneous reproduction number (R_t_) declined over time similarly in 30 largest metropolitan areas, and after April 15, R_t_s in almost all areas reached 1 or below. Since then, the R_t_s remained around 1 in most areas and there were a few small and short rebounds in some regions, suggesting the epidemic was persistent in those areas. The main decline was concurrent with the implementation of aggressive interventions in the US, suggesting stringent interventions were effective in halting the epidemic. However, there were large geographic variations in the R_t_ patterns, partly due to different levels of interventions, geographic altitudes, and partly might be due to temperature variations.

Without effective interventions, the peak of epidemic will reach higher and the epidemic process will last longer [25]. Thus, the reproduction numbers will not decline until a large proportion of susceptible people are infected. For example, during the week of March 25, R_t_s in Houston, Miami, and Orlando were relatively stable (Figure 1a). After the end of March, due to national efforts in mitigating the epidemic, all R_t_ curves started declining. The state of Florida, however, did not officially issue the stay-at-home rule until April 3, 2020, where the curves already declined significantly. This posed some difficulties in assessing the intervention effects precisely. On the other hand, a few cities demonstrated some significant impact of interventions on mitigating the epidemic. For example, some upper midwestern cities (e.g., Minneapolis-St. Paul and Milwaukee) implemented interventions earlier, had lower peak R_t_s, and their R_t_s started declining early, while cities like Pittsburg and Detroit had much higher R_t_ at the beginning, and R_t_ declined later. Even for cities like Chicago and New York, the intervention effects were evident based on the sharp decline of R_t_ since the mid-March, despite they had much higher R_t_s in the beginning of epidemic.

It has been suggested that like many other respiratory virus infections, a seasonal pattern may exist for SARS like coronavirus [26, 27]. However, as demonstrated in this study, the association between reproduction number and temperature was deeply confounded by interventions and other external factors (including altitudes and humidity). In this study, we found the peak R_t_s in warm cities were moderately lower on average than those in cold cities, suggesting that the virus transmissibility might be lower in warm temperature than cold temperature.

However, our analysis warned readers about interpreting selective findings. For emerging epidemic like COVID-19, many things were happening simultaneously. Effective interventions such as travelling restriction, social distancing and stay-at-home rules will change the epidemic process [25, 28, 29]. The availability of testing, diverse case ascertainment criteria, the delay of diagnosis and case isolation, incomplete contact tracing, the percent of asymptomatic cases, and the infectivity of pre-symptomatic and asymptomatic cases will profoundly affect our ability to understand the epidemic. In this study, we observed some small but possible negative association between temperature and virus transmissibility (Figure 1a and 2). However, there were large variations in the peak R_t_s among regions with lower temperature, partly due to different intervention effects and also might be due to cultural and social differences. It is also likely that other environmental factors such as living conditions may affect virus transmission. For example, under cold weather, most people will stay indoors and have close and more frequent contacts with other people. The indoor environment such as air conditioning may be conducive for virus transmission.

Furthermore, the concurrent decline of R_t_ and increase of temperature over time are also confounded by the epidemic process itself. As suggested before, a significant reduction of susceptible people will lead to a decline of R_t_. Longitudinally comparing the trend of R_t_ and temperature is difficult if the epidemic evolves rapidly, as in the current COVID-19 epidemic. For instance, from March to April, all the epidemic in the 30 metropolitan areas had a sharp decline, while the temperature in the whole US was still amenable for virus transmission.

There were some limitations in our study. The most important limitation was the inability to account for the diverse detection capacities across regions (Appendix Table 1). In regions with lower detection capacity, not only were there fewer cases detected (especially missing those with no or mild symptoms), but also the eligibilities for detection were more stringent. Only those with symptoms might be offered for virus detection. Thus, a sudden increase in case counts might not be due to an actual increase of infected people, rather it reflected the increased availability of detection kits. This was the main reason we had highly variable estimates of R_t_ in the beginning of epidemic, and also some small rebounds in some areas after April 15. Additionally, with more detection kits available, we will observe more asymptomatic or mild symptomatic cases. Our methods assumed the same virus infectivity between symptomatic and asymptomatic cases, which was likely not true.

Methodologically, our estimation of R_t_ relied on many assumptions. R_t_ is determined by both the growth rate of new cases and the distribution of generation interval or serial interval [10]. We assumed a universal distribution of serial interval for all regions and over the whole time period. Serial interval may change due to interventions, regional characteristics, and the stage of epidemic. More stringent interventions and stay-at-home rules may result in shorter serial interval because the transmission will likely occur inside the households.

We were not able to rigorously evaluate the virulence of SARS-CoV 2. Although we briefly compared death rates across regions, due to large and unknown delays between virus infection and deaths in the US, most deaths would be diagnosed several weeks before. There was also a delay in death certifications. Additionally, most died cases were elderly people or those with existing chronic conditions. Therefore, assessing the virulence should untangle the confounding effects by health care resource capacities, evolving treatments, and patient’s characteristics. A possible measure of virulence is the pattern of hospitalizations, as they are less likely affected by the availability of detection. After more hospitalization data are available and more asymptomatic and mild symptomatic cases are diagnosed, future research should focus on the impact of epidemic on the severity of disease and health care resource uses instead of the magnitude of epidemic.

In addition, as mentioned before, secondary data analysis based on existing aggregated data suffer many types of unmeasured confounding. Our data were collected by journalists which might lack scientific rigor. However, as stated in the New York Times data website, data were scraped from data portal such as those John Hopkins University COVID-19 data portal, state COVID-19 portal and various press conferences, and verified against each State Health Department. Nonetheless, although the quality of New York Times data was considered acceptable, the data were hastily collected and little validation was done. Biased data might lead to biased conclusions and wrong governmental decisions. We should call for replicated analysis based on more rigorously collected and validated data. Thus, our analyses were mostly descriptive and deliberately avoided over-interpretations through numerous comparisons between regions. A careful exploration of validated individual level data may shed some lights on these issues.

Finally, our study has some unique strengths. First, we focus on large metropolitan areas to ensure enough cases and to have comparable societal structure, individual behaviors and health care resources. Certainly, epidemic in non-metropolitan areas should also be studied and may have unique patterns different from that of metropolitan areas. Epidemic in rural areas was of particular concern recently as rural areas often had limited health care resources to cope with the epidemic. Second, we explored the changes of effective reproduction number (R_t_) over time and across regions to understand the impact of interventions and temperature on virus transmissibility. We presented and compared all regions to avoid selective reporting bias. To study the association between temperature and the epidemic process, we focus on the comparisons of R_t_ at the peak of outbreak and before interventions, presumably less confounded by interventions and other factors.

In summary, we observed a large decline of instantaneous reproduction numbers over time during the COVID-19 epidemic in 30 US metropolitan areas, the timing of which was concurrent with the implementation of mitigating interventions. Given that the whole population were naïve in immunity against this virus, it was likely the epidemic might last longer and more severe without interventions[25]. In addition, there was a possible weak negative association between instantaneous reproduction number and temperature. However, whether this predicts the coronavirus will disappear in the summer and never come back, like that of 2003 SARS coronavirus, is hard to tell. Given that the R_t_s remained around 1 for most areas since later April and the virus is circulating in both northern and southern hemisphere, we need to be vigilant about a possible second wave of outbreak later of the year.

## Data Availability

www.github.com/xinhuayu

## Acknowledgement

We sincerely thank two anonymous reviewers for their insightful comments that significantly improved this report. We also wish to thank Dr. Cheng Zhu from George Institute of Technology for his great suggestions during the study.

## Appendix

**Appendix Table 1:** Positive detection rates at various time by state.

| State | Metro areas | %positive as of March 25, 2020 | %positive as of April 15, 2020 | Date of stay at home rule | % positive at rule mandating |
| --- | --- | --- | --- | --- | --- |
| CA | Los Angeles, St. Francisco-St. Jose, Sacramento-Oakland | 12.9% | 10.6% | 19-Mar-20 | 9.5% |
| CO | Denver | 11.8% | 20.4% | 11-Apr-20 | 19.9% |
| DC | Washington DC - Baltimore | 11.4% | 19.3% | 1-Apr-20 | 15.2% |
| FL | Miami-Fort Lauderdale, Orlando | 9.9% | 10.4% | 3-Apr-20 | 10.5% |
| GA | Atlanta | 20.2% | 23.1% | 2-Apr-20 | 23.3% |
| IL | Chicago | 13.1% | 21.0% | 21-Mar-20 | 12.1% |
| IN | Indianapolis | 14.2% | 18.7% | 24-Mar-20 | 12.5% |
| KS | Kansas | 5.1% | 9.9% | 30-Mar-20 | 7.5% |
| MA | Boston | 9.3% | 22.9% | 24-Mar-20 | 8.4% |
| MI | Detroit | 48.4% | 31.0% | 24-Mar-20 | 52.6% |
| MN | Minneapolis-St. Paul | 2.5% | 4.6% | 27-Mar-20 | 2.8% |
| MO | St. Louis | 49.1% | 10.1% | 3-Apr-20 | 9.8% |
| NC | Durham-Raleigh, Charlotte | 4.8% | 7.7% | 30-Mar-20 | 6.3% |
| NV | Las Vegas | 7.0% | 11.7% | 1-Apr-20 | 10.0% |
| NY | New York | 29.8% | 40.4% | 22-Mar-20 | 24.7% |
| OH | Cleveland, Columbus, Cincinnati | 4.8% | 11.2% | 22-Mar-20 | 71.5% |
| OR | Portland | 4.6% | 5.0% | 23-Mar-20 | 5.0% |
| PA | Philadelphia, Pittsburg | 9.1% | 19.6% | 1-Apr-20 | 12.0% |
| TX | Houston, Dallas-Fort Worth | 7.2% | 10.4% | 2-Apr-20 | 9.2% |
| UT | Salt Lake City | 5.1% | 5.4% | 27-Mar-20 | 5.2% |
| WA | Seattle | 9.4% | 8.7% | 23-Mar-20 | 7.8% |
| WI | Milwaukee | 5.5% | 8.6% | 25-Mar-20 | 5.5% |

**Appendix Figure 1a – d.**
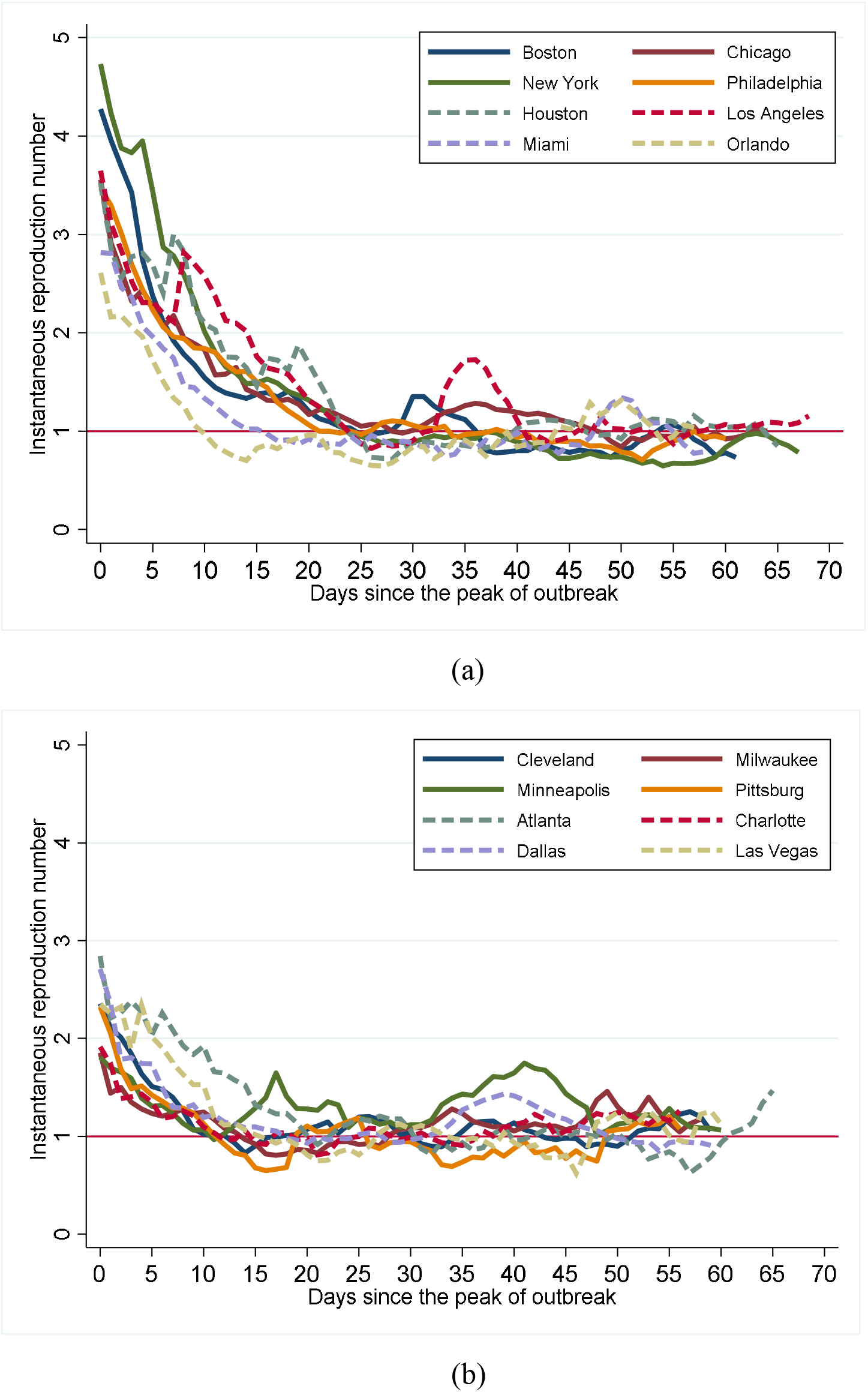
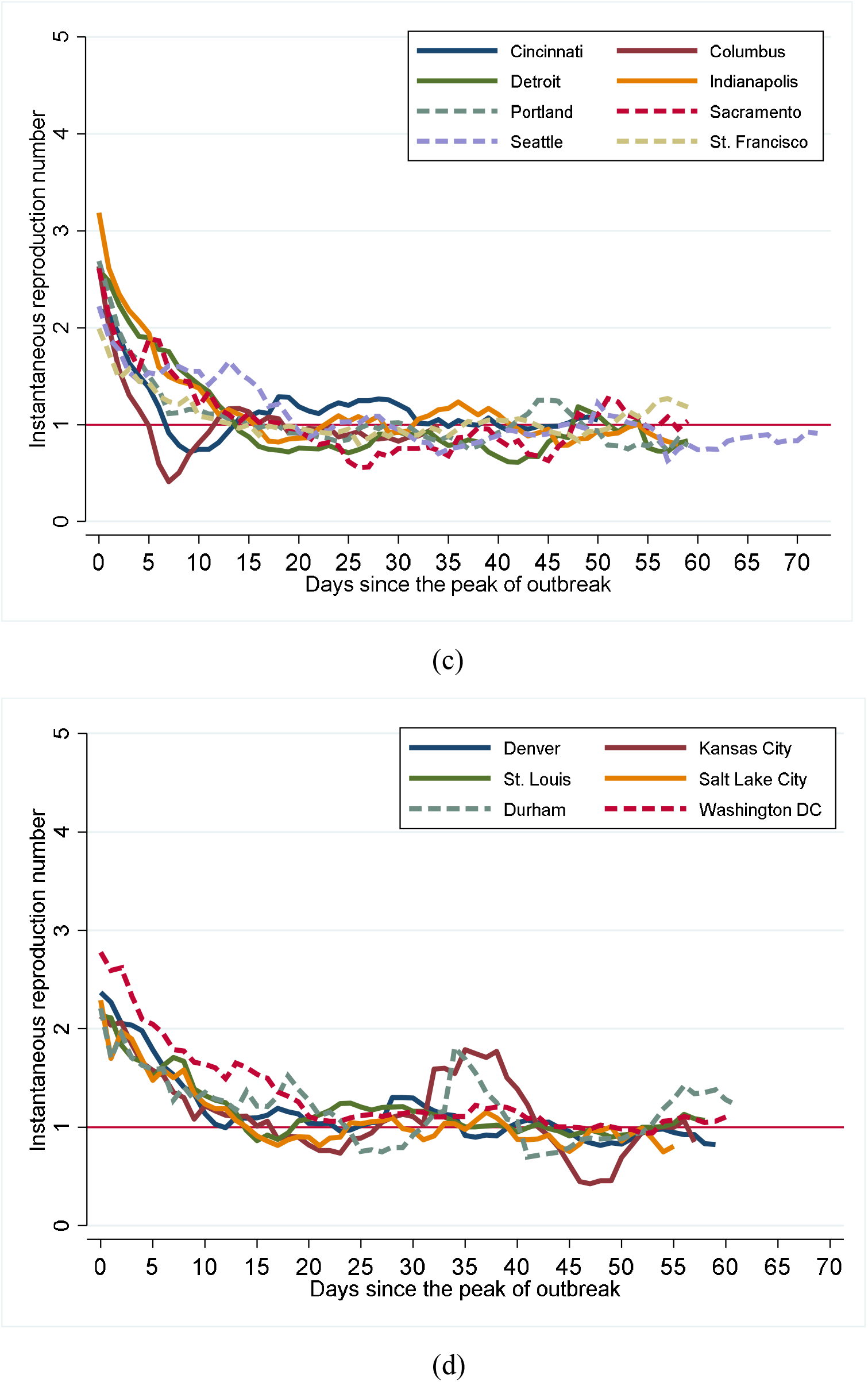
Declining trend of instantaneous reproduction number over time after the peak of epidemic, US 30 metropolitan areas.

**Appendix Figure 2a-b:**
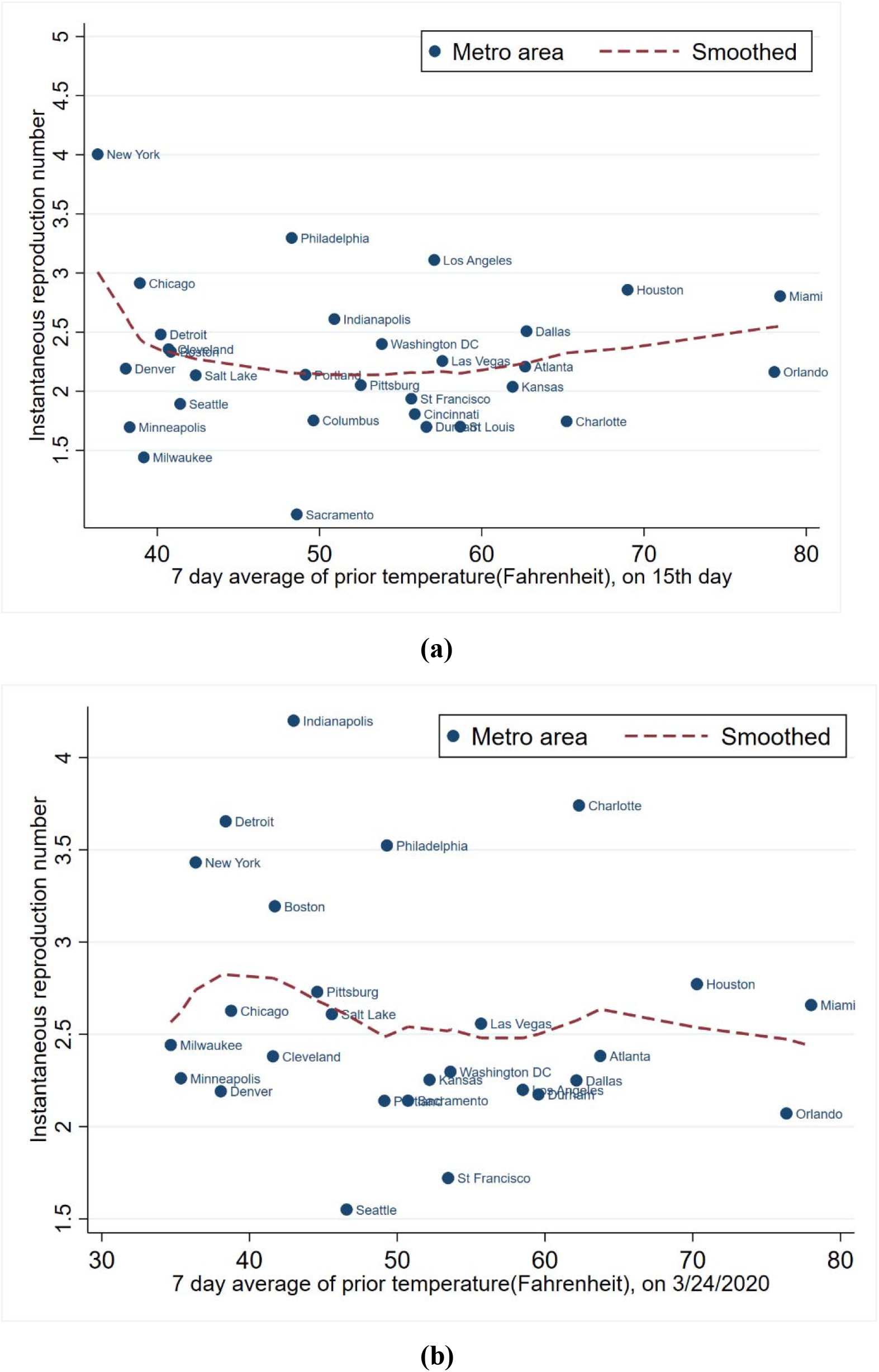
Variations of instantaneous reproduction numbers across 30 US metropolitan areas.

